# *The Feasibility of the Thai Sensory Profile Assessment Tool (TSPA)* for Classifying the participants for Mind-Body intervention

**DOI:** 10.1101/2022.06.05.22275861

**Authors:** Thanaporn Kanchanawong, Thitichaya Prasoetsang, Faungfah Limvongvatana, Ladarat Ooraikul, Sukonta Kunapun, Tiam Srikhamjak, Patama Gomutbutra

## Abstract

**Background:** The previous research found that multisensory interventions impact behavioral responses in healthy individuals and clinical populations, but few have been shown the impacts of Mind-Body intervention on cortisol levels in different sensory personalities. In Thailand, the earlier study found that the Thai Sensory Profile Assessment Tool (TSPA) had acceptable validity and reliability in classifying sensory personality called “sensory pattern”. The pattern is divided into sensory preference and sensory threshold, which is formulated from personal experience.

**Objective:** The main objective of this study is to examine the feasibility and interpretability of TSPA in classifying sensory patterns of participants attending Mind-Body Intervention consisting of Mindfulness-Based Flow Practice (MBFP) and Relax On-site program.

**Materials and methods:** There is a sub-study of a controlled cross-over trial study effect of a mindfulness intervention on anxiety and biomarkers in healthy nursing staff. For this study, we aim to categorize sensory patterns and find the feasibility of Mind-Body Intervention. Each participant was self-tested by TSPA before the intervention, either relaxation on-site or MBFP in the residential retreat program. The feasibility criteria include the time for finishing the test. The interpretability including participant’s comprehension and the classification phenotype provides insight into different effects of the intervention. The effect of MBFP and relaxation on-site was measured quantitatively by the change of morning cortisol before and after the intervention and qualitatively from satisfaction interviews after the intervention.

**Results:** The TSPA assessment takes an average time of 20 minutes. There is no complaint about the difficulty to understand participants. We classified participants by TSPA pattern into three groups by the sensory preference, including, 1) Balanced majority (14/20) have a moderate sensory preference and threshold, 2) low sensory preference for taste and smell (3/20), and 3) high sensory preference for sight smell and movement (3/20). At the same time, most participants show higher cortisol after relaxation on-site and decreased cortisol after MBFP.

**Conclusions:** This pilot study showed that TSPA can be a feasible tool for assessing the sensory preference of the participants to match the health promotion modalities appropriately. We also got a preliminary insight that people with low smell sensory preference, low smell sensory threshold, high sensory preference insight, and movement with moderate threshold showed differences in MBFP. However, it needs a larger sample, and a simpler questionnaire related to the MBFP intervention program to prove these initial findings.

## Introduction

The recent systematic reviews showed personal sensory processing patterns influence quality of life(1) and the effectiveness of mental health intervention(2). Sensory processing refers to a person’s ability to take in, organize and respond to sensory information in the context of the environment(3). The sensation is an objective and simple conscious experience associated with stimuli. Research has shown that each person processes sensory information in different ways (4). For example, some individuals like to work in quiet surroundings, others may like to work while wearing headphones with rap music playing loudly(5). Many studies have investigated relationships between sensory processing styles and behavioral, and emotional responses in all ages either healthy individuals or clinical populations. Elie Chamoun et al demonstrate the application of taste preference and sensitivity/threshold correlates with eating behavior(6). There was also a study that found the effectiveness of mindfulness was significantly related to sensory processing patterns classified by Adolescent/Adult Sensory Profile (AASP). (7)

There were several tools used for classifying the sensory processing patterns(1). The most popular one is Brown and Dunn’s AASP(8) which generates four sensory processing patterns including low registration, sensation seeking, sensory sensitivity, and sensation avoiding. Other modified tools, for example, the Sensory Responsiveness Questionnaire-Intensity Scale (SRQ-IS)(9), and Short Sensory Profile (SSP)(10). However, those tools are generally complicated and interpretation needs clinical expertise.

The Thai Sensory Profile Assessment Tool (TSPA) was developed to be simpler for measuring individual effects of sensory stimuli events in daily life.This tool developed by Tiam Srikamjak et.al. The Thai Sensory Profile Assessment Tool (TSPA). TSPA was designed for measuring individual effects of sensory stimuli events in daily life. The tool is adapted from Dunn’s theory of adult sensory profile(4) and is divided into two parts a) sensory preference which is quite stable formulated from personal experience, and b) sensory threshold, which is more labile, that determines the intensity of sensory perception. A person with a high sensory threshold would perceive less intensity than those who have a low threshold. The earlier study found that TSPA has acceptable validity and reliability to classify sensory processing patterns(11). Although further verification of this version of TSPA is needed, benefits from its applications for Thai people have been shown. For example, Kongngern and Srikamjak used TSPA to investigate the relationship between Sensory Patterns and Stress in 90 relapsed alcohol-dependent clients in Suansaranrom Psychiatric Hospital, Surat Thani province, Thailand. The results of the study portrayed that: 1) sensory preferences of vision had negative significant correlation with stresses (r = - .264, .341, P < .05) and was the most influence on Stress (Beta = - .433, .511, P < .05), 2) sensory threshold of smell had negative significant correlation with stress (r = - .245, - .263, P < .05) and was the most influence on stress (Beta = - .349, - .292, P < .05)(12). The Psychometric properties of TSPA have been examined. Content validity was determined in conjunction with an expert panel discussion. The content of the TSPA was accepted by a majority of the experts, psychiatrists, occupational therapists, physical therapists, psychologists, and special education teachers. The reliabilities of the tools were assessed using data from 387 Thai participants aged 15 years and over from 5 geographical areas of Thailand. The test-retest reliability results of Modules 1 and 2 were obtained using the Intraclass Correlation Coefficient method. These were between 0.78 - 0.85 for Module 1 and between 0.55 - 0.75, for Module 2. The internal consistency, using Cronbach’s correlation coefficient, for Module 1 was between 0.59 - 0.78, and for Module 2 was between 0.32 - 0.62. (13)

Therefore, the purpose of this pilot study was to examine the feasibility and interpretability of TSPA for classifying sensory patterns of participants who attend the Mindfulness-Based Flow Practice (MBFP) and Relax On-site program. The feasibility criteria include the time for finishing the test. The interpretability including participant’s comprehension and the classification phenotype provides insight into different effects of the intervention. The effect of MBFP or relaxation on-site was measured quantitatively by the change of serum morning cortisol before and after the intervention and qualitatively from satisfaction interviews after the intervention.

## Materials and methods

### Participants

Our target population is healthy people with a high risk of anxiety and burnout. Based on epidemiologic studies, working females, especially health personnel is one of the highest prevalence of burnout(14). Therefore the inclusion criteria of this study were physical healthy nursing staff working in a university hospital who has the Generalized Anxiety Disorder: GAD-7 showed mild anxiety (score 5 or more) (15) or the Depression score: PHQ-9 showed depression (score 7 or more) (16). Participants were recruited from the self-enrolled volunteers from the palliative care nursing Department in the Faculty of medicine. The participants consisted of nurses and nursing assistants. All participants have informed of the purposes of the project and the extent of their involvement and gave their consent before participating in the study.

### Procedures

This study was approved by the Faculty of Medicine, Chiang Mai University, Thailand, as a sub-study under the project entitled “The Cellular and Physiologic Mechanism of Mindfulness-Based Flow Practice (MBFP) therapy for Depressive and Anxiety Symptoms, an Exploratory Clinical Trial”, and by the Faculty of Medicine, Chiang Mai University, Institutional Review Board (IRB), the Chiang Mai Medical School Ethical Committee (study code: FAM-2561-05432). Participants were informed consent before data collection. Participants were randomized into two groups, there are group A and group B. Group A provided the intervention (MBFP) in the first period and Relax On-site program in the second period. On the other hand, For group B received the Relax On-site program in the first period, and in the later period group, B participated in the intervention(MBFP). The first and second periods took three months for the wash-out period. Besides that, the researcher made an appointment with the technician to take the blood of the participants for checking serum cortisol in the morning. One day before and after the first and second periods (Friday and Monday) in each period. On the day of the first period, the researcher collected TSPA data from both groups in the morning before attending the Mindfulness-Based Flow Practice (MBFP) and Relax On-site program. NCSS version 2021 (East Kaysville, Utah) was used for analysis. The demographic data were described in frequency and mean. The sensory patterns data were analyzed in frequency, percentage, and quartile deviation. Furthermore, after attending the intervention in the first and second periods the researcher collected the qualitative data by using the interview with participants about their satisfaction with the Mindfulness-Based Flow Practice (MBFP).

### Intervention

#### Mindfulness-Based Flow Practice (MBFP) and Relax On-site program

In both the Mindfulness-Based Flow Practice (MBFP) Group and Relax Onsite group, all participants underwent 8-hour on weekends (Saturday and Sunday)

1. Two Hours Mindfulness-Based Flow Practice (MBFP) 2- hours MBFP is a mind-body intervention integrated the concepts of flow and mindfulness model through 6 activity modules including 1) 10-15 minutes of learning about MBFP (before or after practices) 2) 10-15 minutes of self-observation through the sensory, emotional, and cognition information 3) 40-45 minutes of muscle stretching 5) 15 minutes of relaxation breathing 6) 10-15 minutes of imagery guidance. Most activity modules focus on telling participants about their body position or proprioceptive sense.
2. Relax On-site program Participants were assigned to the same resort as the MBFP group and had the same food. During the MBFP Intervention, they were allowed to travel or light exercise around the resort but not allowed to bring office work to do. This program is modified from concepts of multisensory environments / MSEs to promote enjoyment and relaxation.

### Instruments

#### 1. The Thai Sensory Profile Assessment Tool (TSPA): Version 2007

TSPA is a self-administered measurement of sensory responses for the general public over 15 years old who can read and write Thai. The tool is divided into two modules. The first module, sensory preference, is composed of 60 items or questions related to sensory systems such as sight, hearing, smell, taste, touch, and movement which a subject usually likes to use, especially in everyday life. The second module, sensory threshold, is composed of 60 items or questions related to sensory systems including sight, hearing, smell, taste, touch, and movement which a subject usually needs for responding to stimuli in everyday life. The TSPA measures the respondent’s frequency of responses to specific sensations by employing a Likert scale from 0 = never to 4 = always for sensory preference and high threshold questions, and from 4 = never to 0 = always for low threshold questionnaires. Scores from both modules are calculated and reported as graphs and percentages by a computer-based program as shown in Figure 1.

**Figure 1:**
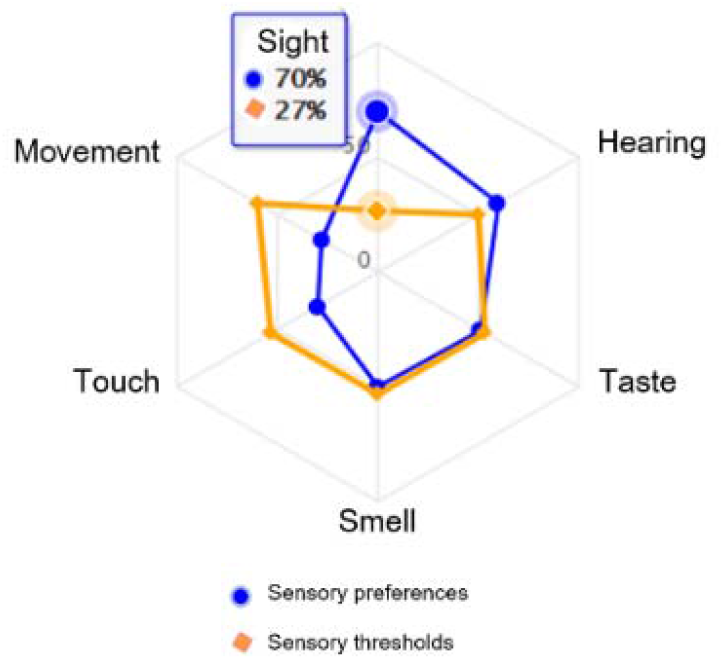
TSPA Report: blue line show the sensory preference and the orange line is the sensory threshold

The interpretation of the TSPA report is the cut scores. The cut scores do not indicate at which point a particular pattern becomes problematic but, instead, show a particular frequency and individual response to sensory stimuli in everyday life. This tool uses percentage and quartile deviation of each sense as cut scores consist of 1) Less than 25 % = Low score 2) 25-75% = Moderate score and 3) More than 75% = High score.

#### 2. Morning serum cortisol

Morning serum cortisol serum is a common physiology biomarker that may be used for examining the biomarker outcomes of the Mind-Body Intervention (MBIs). For this study, blood samples were collected between 8-9 am and stored in EDTA-coated tubes in the morning. Plasma was obtained by centrifugation of blood at 3000 rpm, 4 °C 10 minutes, and frozen at -80 °C immediately until analysis. Plasma cortisol levels were measured and analyzed according to the protocols provided by the test manufacturer at the central laboratory of the Maharaj Hospital, Faculty of Medicine, Chiang Mai University, Chiang Mai, Thailand.

## Results

### Demographic data

The participants in the pilot were 20 palliative care nursing staff, who were working in a university hospital, aged 20-45 years old. All participants were healthy and did not have an uncontrolled disease; 75% are single. The mean Generalized anxiety disorder score was 6.89. None of the participants has a Depression score (PHQ-9) of 7 or above and the mean score is 4.78. The mean morning serum cortisol level both before and after was 10.19 μg/dl.

### The feasibility

The satisfaction shows feasibility criteria categorized by choosing the highest sensory preference and threshold of each score.

#### 1. Dominant characteristic in Sight part

The example interview data from participants who had a high score of sensory preference in sight was outlined below.

· *“The dark light made them focus.”*
· *“The light made me concentrate on the program.”*
· *“The light makes me comfy and cozy.”*
· *“The dim light made me feel calm and relaxed.”*
· *“The light in this program creates the relaxed vibe.”*
· *“The dark light made me improve my focusing on myself and the intervention.”*

#### 2. Dominant characteristic in Sound part

The satisfaction comments from participants review the feeling of music in this program. For instance,

· *“The music made me feel relieved”*
· *“The music helps me focus on the present moment”*
· *“The sound that was used in this program was so comfortable and not too exciting for me.”*
· *“The description during the flow program instructor made me feel the flow.”*
· *“The sound of rain and waterfall help me calm and easily fall asleep.”*

#### 3. Dominant characteristic in Touch part

Moreover, participants with dominant scores in touch sensory gave interview data below.

· *“I like the hugging position from this program it’s made me feel warm every time I hug myself”*
· *“The massage part is good and restful.”*
· *“The message from this program helped release my muscle pain.”*
· *“I like the massage part of this program.”*
· *“I need more massage in this program”*

#### 4. Dominant in Movement part

There is interesting data from pilot cases which is divided into two parts consisting of preference and threshold.

##### 4.1 Threshold

· *“This program promotes body movement and helps me relax.”*
· *“It’s made me feel like I’m exercising but it is not hard. After this program, I feel so comfortable.”*
· *“I need to work hard to follow the instructions of the instructor to stretch my body. Lastly, I can do it and I feel better and calm.”*

##### 4.2 Preference

· *“This intervention helps stretch the muscles that rarely move for a long time*.
· *“I feel relaxed and have happy energy after this intervention. “*
· *“The stretching phase in this intervention releases the pain like I am exercising.”*
· *“I like stretching my muscles.”*
· *“The tenses of the muscles make me feel like doing exercise.”*

Furthermore, there was positive feedback from high-score participants below.

> *“It makes me relaxed and I feel the substance of happiness in my body increasing” (High preference for movement)*
>
> *“dark room makes have better sleep and it gives me nice resting time” (High threshold of sight)*
>
> *“It seems like exercise for me, but it makes me feel relieved”*
>
> *“The sound makes me feel peaceful”*

***The interpretability and preliminary insight***.

The qualitative data from the interview

Overall, most of the feedback was positive as follows.

> *“I like self-hug posture, it makes me feel warm”*
>
> *“Stretching muscle makes me feel relaxed”*
>
> *“Feel comfortable and peaceful”*
>
> *“Massage relieves my muscle pain”*
>
> *“It’s nice to rest for me”*

However, there was some negative feedback to the movement section from participants with the dominant characteristic of movement sensory preference as follows.

> *“I didn’t like the difficult postures because I could not do them.”*
>
> *“Some postures gave me pain (especially trunk hyperextension.”*

The massage section (that could be represented as the tactile section) from participants with dominant characteristic of tactile sensory preference.

> *“I think the amount of compression was inadequate for me.”*
>
> *“The massage tickled me, so I didn’t like it.”*

The visual part from participants with tactile sensory preference.

> *“Sometimes, I felt distracted from the light outside the room.”*

The sound background from participants with dominant characteristic movement and touch sensory preference

> *“I didn’t like it when there were several sensory stimulations because I prefer to be in a quiet place during leisure time.”*
>
> *“Sometimes, the sounds were scary to me.”*

Most of the participants had a moderate sensory preference (see Table 2). One participant scored a low sensory preference in taste and 2 participants in smell. Three participants had high sensory preferences in sight, smell and movement.

**Table1:** Demographic data and measurement outcome

|  |  |  |
| --- | --- | --- |
| Factors |  | (N=20) |
| Female sex | n (%) | 20 (100) |
| Age (year) | Mean ± SD | 28.0 ± 12.55 |
| Occupation |  |  |
| Registered nurse | n (%) | 18 (90) |
| Nursing assistant | n (%) | 2 (10) |
| Education |  |  |
| Master | n (%) | 4 (20) |
| Bachelor | n (%) | 12 (60) |
| High school | n (%) | 4 (20) |
| Marital status |  |  |
| Single | n (%) | 15 (75) |
| Married | n (%) | 5 (25) |
| Generalized anxiety disorder score (GAD -7 score) | Mean ± SD | 6.89 ± 2.99 |
| Depression score (PHQ-9) | Mean ± SD | 4.78 ± 3.41 |
| Morning serum cortisol (µg/dl) | Mean ± SD | 10.19 ± 5.59 |

**Table 2:**
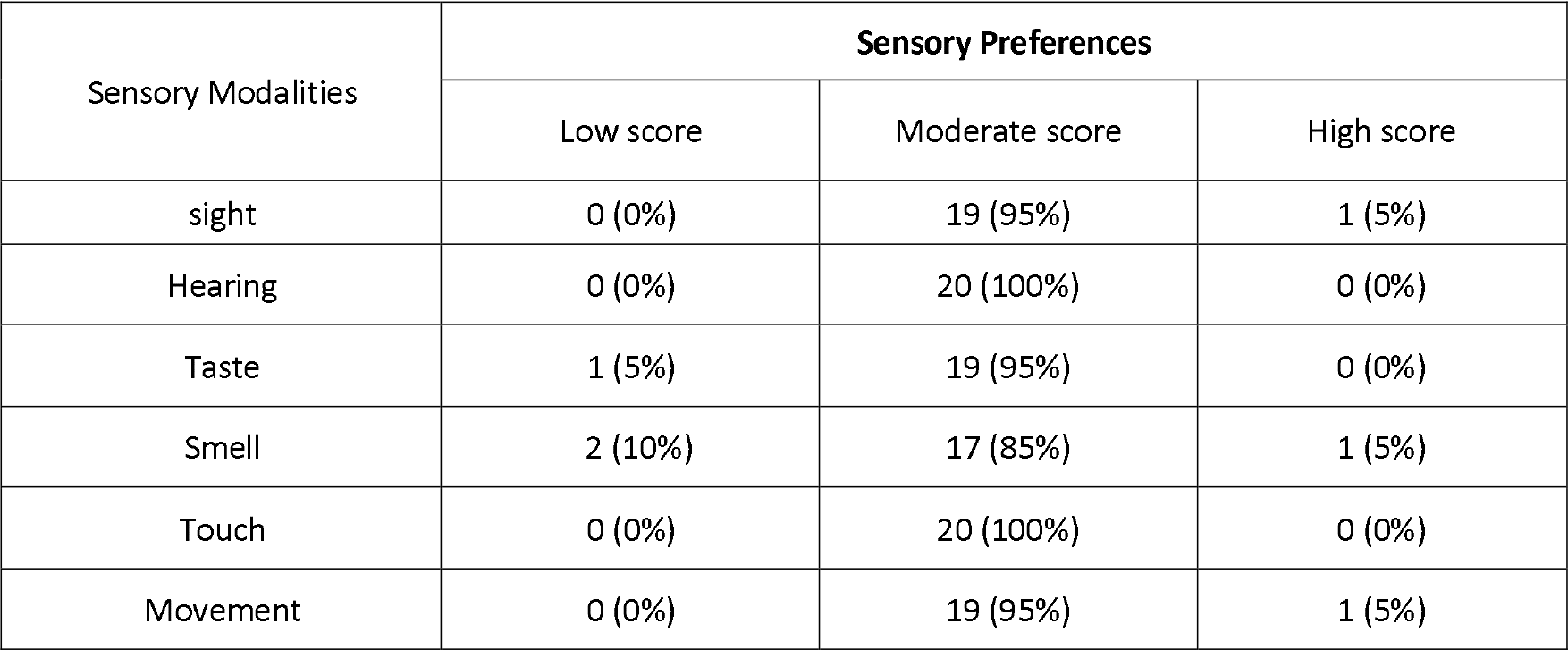
Frequency and Percentage of participants’ Sensory Preferences clarified by TSPA (N=20)

The sensory threshold for each group of sensory preferences is shown in Table 3. It could be noted that all the participants who had moderate sensory preference also had a moderate sensory threshold. Among the three who had low smell preference, two had low smell preference, but one had a high smell and sight threshold. The high sensory preference group all has moderate sensory thresholds.

**Table3:** Frequency and Percentage of participants’ sensory thresholds clarified by TSPA for each group of sensory preference (N=20)

| Sensory Modalities | Sensory Thresholds |  |  |
| --- | --- | --- | --- |
|  | Low score | Moderate score | High score |
| Moderate all sensory preference | 0 (0%) | 14 (100%) | 0 (0%) |
| Low smell preference | 2 (10%) taste, smell | 0(0%) | 1 (0%) sight, smell |
| High smell preference | 0(0%) | 1 (100%) all sense | 0 (0%) |
| High sight preference | 0 (0%) | 1 (100%) all sense | 0 (0%) |
| High movement preference | 0 (0%) | 1 (100%) all sense | 0 (0%) |

Figure 2 shows the group of participants who had moderate levels in all sensory processing patterns and thresholds showed a similar response pattern, one in which serum cortisol increased after relaxation on-site and reduced after MBFP. In Figure 3, it can be noted that low smell sensory preference with a high threshold demonstrates the same response as the first group. Still, those with low sensory threshold showed blunted responses to both relax on-site and MBFP. Figure 4 shows the group of high sensory preferences, including those with high movement, sight, and smell with moderate in all sensory thresholds. Those with high smell sensory preference demonstrate cortisol change after interventions, the same as the first group. However, those with high sight preference reduce cortisol after relaxation on-site, and those who have high movement preference show a ‘reverse’ pattern contrast with the first group.

**Figure 2:**
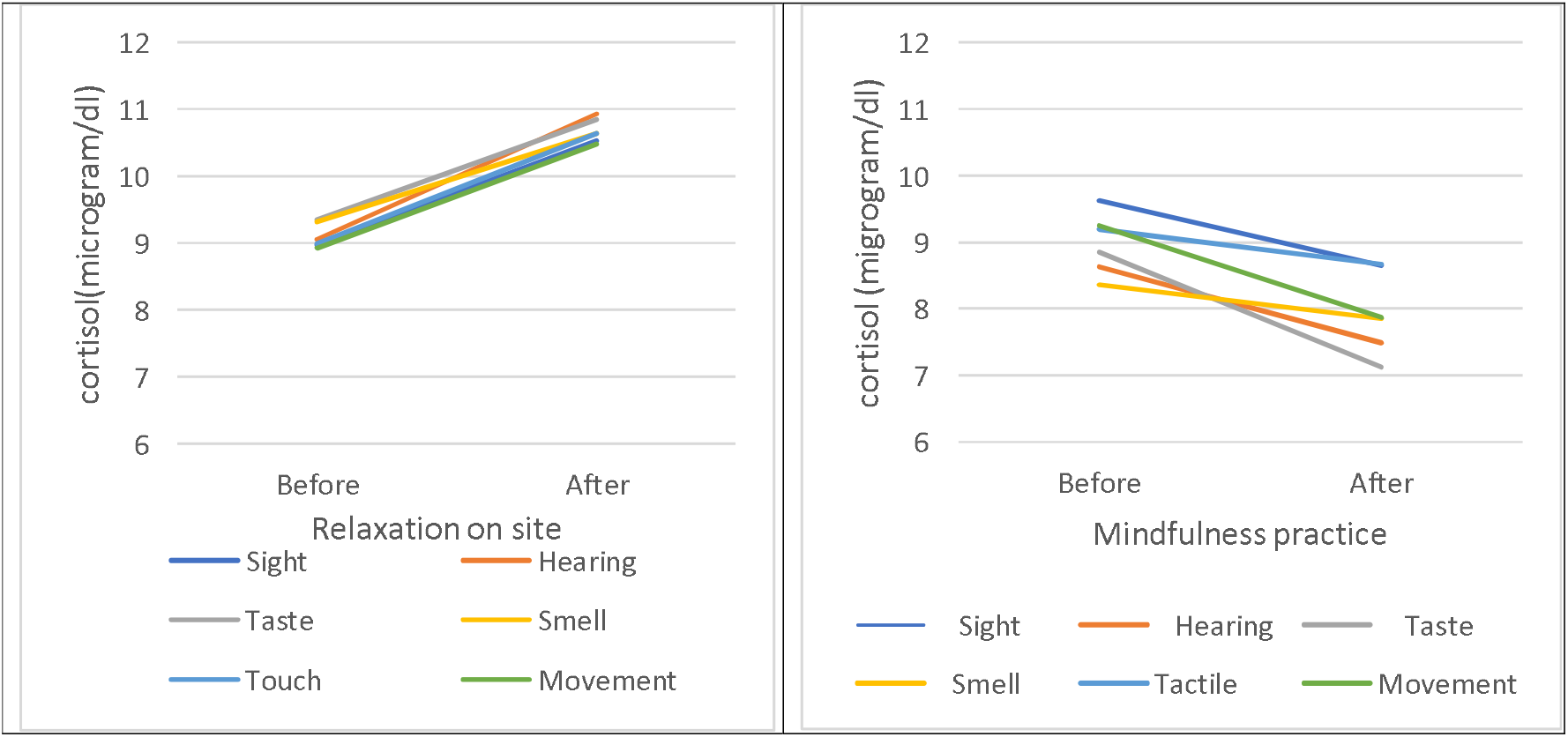
Effects of Relaxation on Site and Intensive MBFP on cortisol levels in participants with moderate scores of all sensory preferences and sensory thresholds group (n = 14)

**Figure 3:**
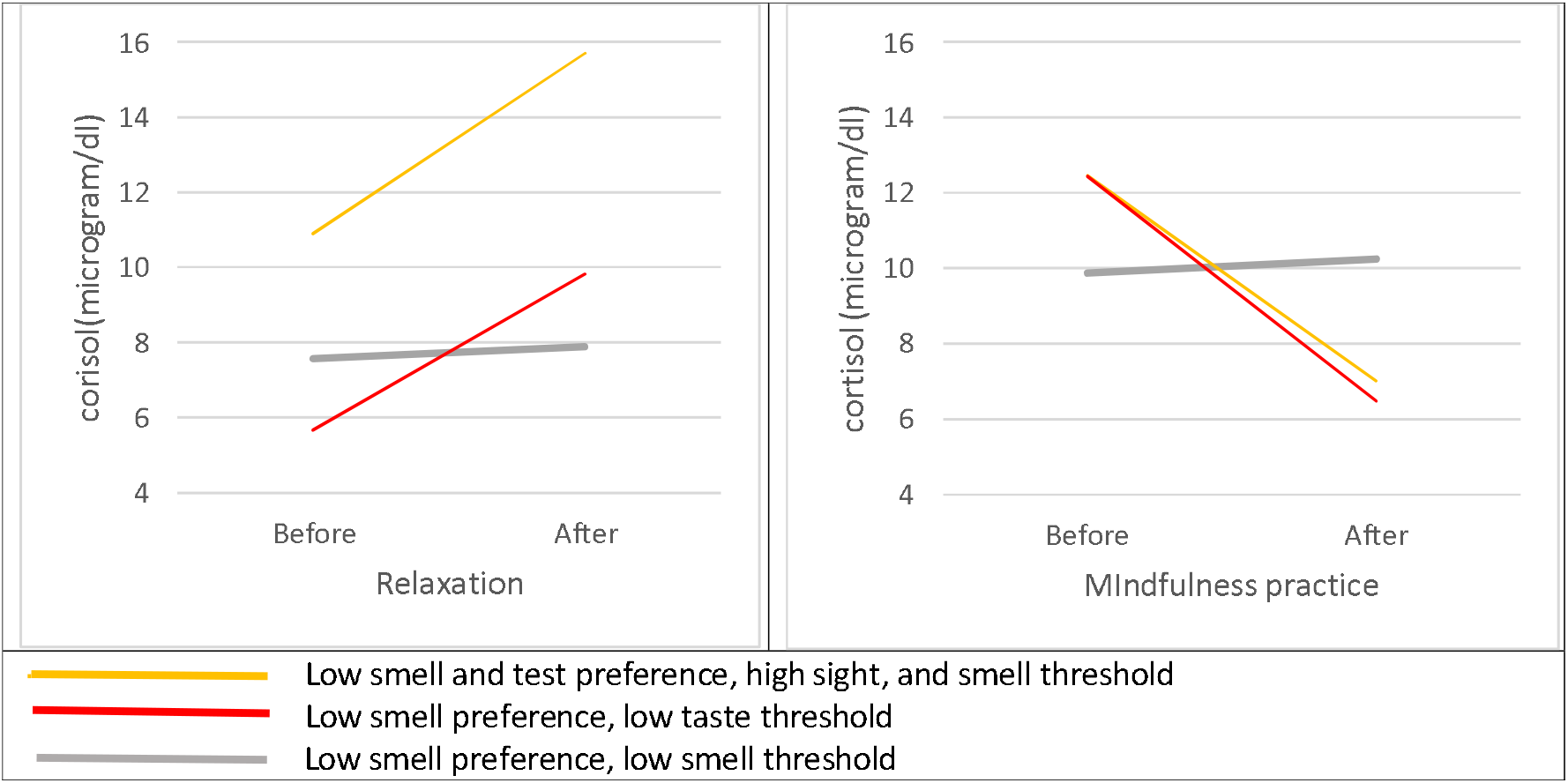
Effects of Relaxation on Site and Intensive MBFP on cortisol levels in participants with low sensory preference in smell and test (n=3)

**Figure 4:**
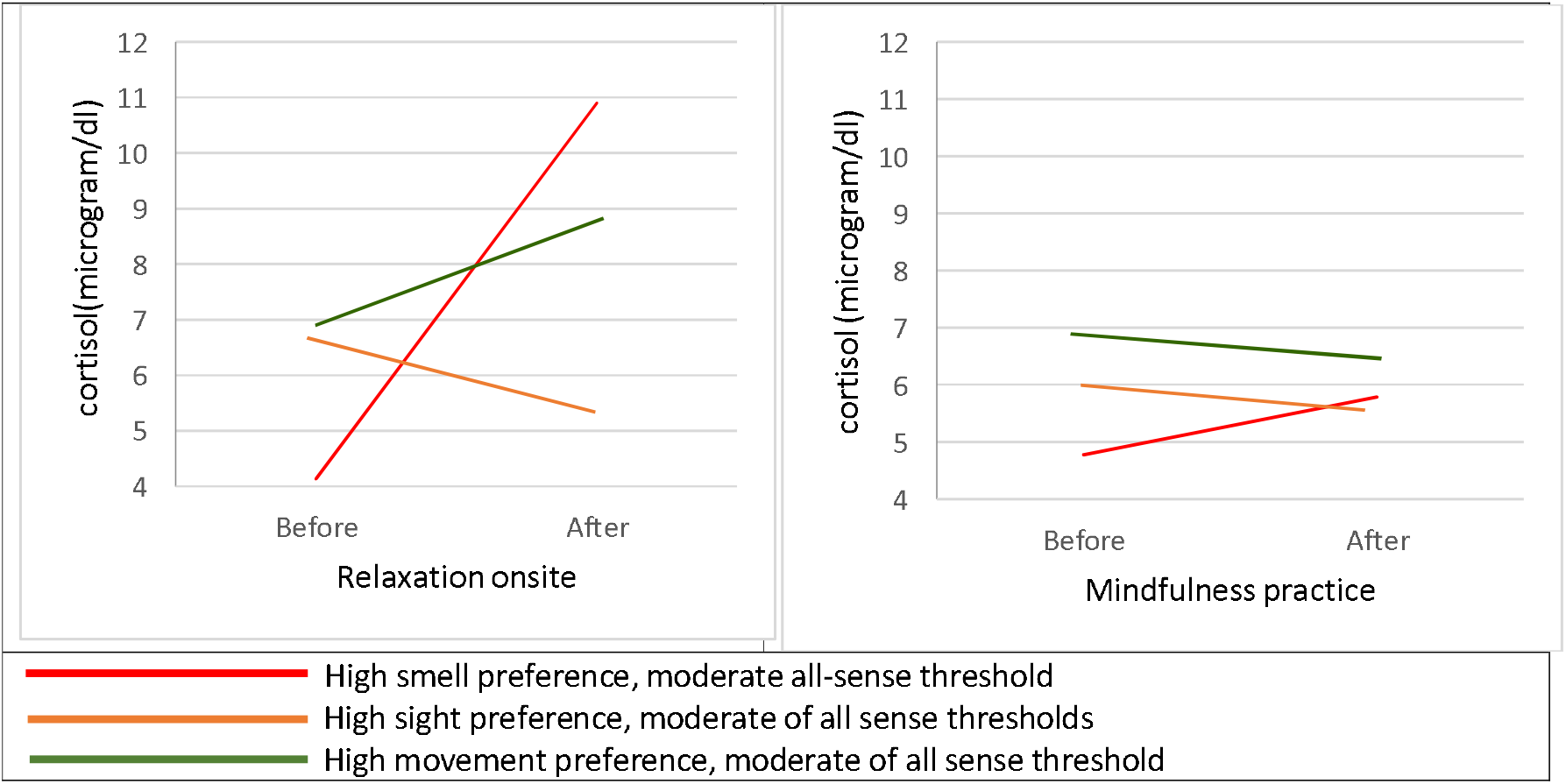
Effects of Relaxation on Site and Intensive MBFP on Cortisol Levels in high preference of smell, sight, and movement and a moderate score of other senses both preferences and thresholds (n=3)

## Discussion

Our results indicate that TSPA has acceptable feasibility and interpretability used to classify sensory patterns of participants who attend the Mindfulness-Based Flow Practice (MBFP) and Relax On-site program. The feasibility evaluation from the average time used to complete the test is not more than 20 minutes of self-testing and not over 5 minutes for the interview. The interpretability is also acceptable by both qualitative interviews that showed participants’ valid comprehension and the classification provide some initial insight into different cortisol responses to the interventions. It appears that the majority (80%) of participants balance all sensory preferences and sensory thresholds. These results are consistent with a previous study that this sensory pattern is the commonest in the general population. (17). This group responds to the same pattern; increases cortisol after relaxation on-site and reduces cortisol after MBFP. It could be explained by the nature of relaxation on-site, though inhibiting bringing work to do in the report, could not prevent participants from rumination or worrying about their work and family. Meanwhile, MBFP has more potential to bring the participant’s attention to the activity. However, this study could not claim that MBFP reduces cortisol more than relaxation on site. In addition, the cross-over control trial which consisted of 50 participants found that there was no significant difference in cortisol, but MBFP significantly increased Brain-Derived Neurotrophic Factor compared to relaxation on site. (10).

Interestingly, a person with low smell sensory preference and low threshold showed blunted responses to both relaxation on-site and MBFP. This person may be classified as a kind of ‘sensory avoidance’ according to Dunn’s Model of Sensory Processing. Although there are no particular aroma sensory stimuli in either relaxation on-site or MBFP, this personality shows blunted response which is different from the peer. This person also has a high score of burnout (19) which is coherent with a previous study’s finding that those who have sensory avoidance have high levels of working-related burnout(3). In addition, those who have high movement preference and moderate all sensory thresholds show a ‘reverse’ pattern contrast with the first group. It could reflect the nature of the intervention since relaxation on-site allows participants to walk or run around the resort while MBFP limited movements to only joint position sensation (proprioception). This is reflected in an interview with this person *“I didn’t like the difficult postures because I could not do them.”* and *“Some postures gave me pain (especially trunk hyperextension.”*

However, the small sample size also impacted the limited number in each classified sensory pattern, meaning that these results should be interpreted cautiously as Winnie Dunn stated that “the response to sensory stimuli can be more easily understood when we study people whose sensory processing is so unusual that it interferes with even desirable life activities. The concepts involved are not as clear when studying people without a particular disorder because most people have moderate behaviors that don’t stand out”(7).

Usability of TSPA was scored as good within the sub-group of study participants. The study has generated several suggested areas for improvement which could use TSPA as an assessment tool in clinical populations as follows: First, the TSPA consists of 2 modules, sensory preferences, and sensory thresholds. Each module comprises 6 sensory modalities: sight, sound, smell, taste, touch, and movement. The contents of the TSPA were accepted by a majority of the experts. The test-retest reliability results of Modules 1 and 2 were obtained using the Intraclass Correlation Coefficient method. There were between 0.78 - 0.85 and between 0.55 - 0.75 for Modules 1 and 2, respectively. The internal consistency, using the Cronbach’s correlation coefficient, of the Module 1 was between 0.59 - 0.78, and for Module 2 was between 0.32 - 0.62. The movement had the lowest Cronbach’s correlation coefficient which might be because this sensory modality consists of 2 sensory systems: vestibular and proprioceptive sensation. Therefore, the items in this sensory modality might not result in the same measurement, leading to a low internal consistency. For further studies, separation of these two senses, as well as research used in development and refinement, are recommended to maximize reliability and validity. Second, TSPA is divided into sensory preference, which is composed of 60 items, and sensory threshold, which is composed of 60 items. It might have too many items and take too long for the participants to complete. Therefore, we may develop a shorter version of TSPA or even design TSPA to be more suitable with MBFP intervention such as items asking about prefered posture or stimuli according to individual sensory preferences and sensory threshold. Third, technological advancement in the area of mass communication. The trend of the word has shifted everything done on the internet, such as education, entertainment and health care. Incorporate to the widespread of the coronavirus pandemic (covid-19) impact on access to services and resources, and also press on people to change their routine and improve their resilience and self-management(20). Future feasibility of TSPA will need collaborative assessment between individuals, families, therapists, and researchers to better understand individual preferences and thresholds through the activity called “Telehealth”.

## Conclusion

This pilot study showed that TSPA can be a feasible tool for assessing the sensory preference of the participants to match the health promotion modalities appropriately. We also got a preliminary insight that people with low smell sensory preference, low smell sensory threshold, high sensory preference insight, and movement with moderate threshold showed differences in MBFP. In a bigger study, there should be a discussion about participants’ sensory preferences and sensory threshold for designing MBFP programs in detail. However, it needs a larger sample, a shorter version of TSPA, a more specific questionnaire related to the MBFP intervention program, and more intense or alternative sensory stimulation in MBFP intervention to prove these initial findings.

## Data Availability

All data produced in the present study are available upon reasonable request to the authors

## Acknowledgments

This study received kindly support from the Faculty of Medicine and Faculty of Associated Medical Science, Chiang Mai University. Moreover, the researchers thank Professor Dr. Buncha Ooraikul and Professor Dr. Magaret Fitch for valuable suggestions

## Supplement data

The Thai Sensory Profile Assessment Tool (TSPA) could be accessed via DOI 10.17605/OSF.IO/4WV9A for the English version and DOI 10.17605/OSF.IO/498M2 for the Thai version.

